# A survey of the psychological status of primary school students who were quarantined at home during the coronavirus disease 2019 epidemic in Hangzhou China

**DOI:** 10.1101/2020.05.28.20115311

**Authors:** Yanghao Zheng, Jianhua Li, Maiyan Zhang, Bicheng Jin, Xiaoyi Li, Zhiyong Cao, Nanping Wu, Changzhong Jin

**Author notes:** Correspondence to: Dr. Zhiyong Cao Postal address: Greentown Yuhua Primary School of Hangzhou, West Wenyi Road 532, Hangzhou 310012, Zhejiang Province, China., Dr. Nanping Wu Postal address: State Key Laboratory for Diagnosis and Treatment of Infectious Diseases, National Clinical Research Center for Infectious Disease, National Medical Center for Infectious Disease, Collaborative Innovation Center for Diagnosis and Treatment of Infectious Diseases, The First Affiliated Hospital, School of Medicine, Zhejiang University, Qingchun Road 79, Hangzhou 310003, Zhejiang Province, China., Dr. Changzhong Jin Postal address: State Key Laboratory for Diagnosis and Treatment of Infectious Diseases, National Clinical Research Center for Infectious Disease, National Medical Center for Infectious Disease, Collaborative Innovation Center for Diagnosis and Treatment of Infectious Diseases, The First Affiliated Hospital, School of Medicine, Zhejiang University, Qingchun Road 79, Hangzhou 310003, Zhejiang Province, China.

## Abstract

**Objective:** To investigate the presence of social anxiety and depression and the risk factors for them among primary school students who were quarantined at home during the coronavirus disease 2019 (COVID-19) epidemic in Hangzhou China.

**Methods:** A total of 1620 students who were quarantined at home for at least one month were recruited from two primary schools in Hangzhou. Students completed a questionnaire on a mobile App with help from their guardians; the measures included demographic and general information, the Social Anxiety Scale for Children (SASC), and the Depression Self-rating Scale for Children (DSRSC).

**Results:** The mean SASC score of the participants was 3.90 ± 3.73, which was higher than the mean norm score of Chinese urban children (3.48 ± 3.47) (P < 0.01). The mean DSRSC score of the participants (5.67 ± 4.97) was much lower than the mean norm score of Chinese urban children (9.84 ± 4.73) (P < 0.05). A total of 279 (17.2%) students had social anxiety, with a mean score of 10.41 ± 2.59, and 102 (6.3%) students had depression, with a mean score of 18.96 ± 3.89. The following variables were found to be significant risk factors for social anxiety during home quarantine: deterioration of the parent-child relationship, increased conflicts with parents, irregular work and rest, and worrying more about being infected. Deterioration of the parent-child relationship, less physical activity, irregular work and rest, and negative mood during home quarantine were significant risk factors for depression.

**Conclusion:** Primary school students who were quarantined at home during the COVID-19 epidemic were more likely to have social anxiety but less likely to have depressive symptoms. Poor parent-child relationships, irregularity of work and rest, and epidemic-related problems were the main reasons for psychological problems. Families, schools, and social organizations need to pay more attention to the psychological status of primary school students quarantined at home.

## Introduction

A new coronavirus began to spread in Wuhan China in mid-December 2019, and then quickly spread all over the country [1]. The novel coronavirus was named “severe acute respiratory syndrome” coronavirus 2 (SARS-CoV 2) and the disease was named coronavirus disease 2019 (COVID–19) [2]. The Chinese government adopted a series of measures to control the epidemic and prevent its further spread, including blocking access to cities, closing schools, and quarantining people at home in the affected areas [3]. On one hand, these measures greatly shortened the duration of the epidemic, and reduced the spread of the virus and the incidence of COVID-19[4]. On the other hand, people’s social activities were restricted by these measures, especially the strict home quarantine measures, which greatly reduced people’s spatial activity. A recent study by Kandola et al. [5] found that sedentary behavior, a decrease in physical activity, was associated with an increased risk of depressive symptoms in adolescents. Children are in a developmental stage with respect to their body and mind and their psychological status is immature and less well understood [6]. Reduced activity in space, insufficient outdoor activities, and especially, reduced contact with peers or classmates may have negative effects on their psychological well-being.

Childhood is a critical period of neurodevelopment, and many mental-health problems occur during this time [7], of which, social anxiety and depressive disorders are the two most common mental-health disorders in children and adolescents [8]. Social anxiety is characterized by emotional discomfort, fear, anxiety, and worrying about social situations [9]. This kind of emotion may promote irrational behavior in teenagers to different degrees, reduce happiness with life, and increase social conflict and dissatisfaction. Depressive disorder is a widespread chronic medical disease that affects thoughts, emotions, and physical health to varying degrees [10]. Depressive disorder is often manifest as low mood, slow thinking, decreased activity, and impaired cognitive function. It has been reported that about 6.5% of minors are in a state of anxiety and 2.6% are in a state of depression [11]. The onset of these mental diseases is relatively slow and insidious, but their consequences are quite serious, ranging from interruption of interpersonal relationships and dropping out of school, to lifelong mental illness and suicidal behavior [12]. Very few psychological problems of adolescents are recognized by their guardians. Even in resource-intensive environments, less than one-third of children and young people in need of mental-health care obtain treatment [13]. Early detection of anxiety and depression in children and adolescents is essential to prevent and treat these mental problems.

An epidemic of serious infectious diseases can have a negative impact on mental and psychological health, such as panic disorder and anxiety [14], and stress disorder may occur in severe cases [15]. The rapid spread of the epidemic and the lack of vaccines and special drugs during the early stage of the outbreak of COVID-19 may have had negative effects on people’s psychology. Strict home quarantine for more than a month could exacerbate this adverse effect because children and adolescents had less opportunity to have contact with their peers and classmates due to school closures and being required to stay at home for a long time. Therefore, their mental status has become the focus of our attention. This study used a survey questionnaire to investigate the psychological state of children quarantined at home during COVID-19, especially social anxiety and depression, and possible factors to determine psychological problems of children in the early stage of the epidemic and provide timely preventive and intervention measures.

## Methods

### Participants

A cross-sectional survey was conducted of all students from grade 1 to grade 6 enrolled in two primary schools (one private and one public) in the urban area of Hangzhou, China, who were under strict home quarantine from January to February 2020

### Measures

#### General Information Questionnaire (GIQ)

The authors developed the GIQ to collect information about demographic characteristics (including school, class, name, sex, age), the parent-child relationship and family support, learning, exercise, and entertainment during the home-quarantine period, knowledge of COVID-19, and emotional changes during the epidemic. Detailed results are shown in Supplemental Table 1.

#### Social Anxiety Scale for Children (SASC) [16]

The SASC, which is a screening tool for symptoms of social anxiety in children developed by La Greca, is suitable for ages 7–16. The scale contains 10 items that measure two dimensions: (1) fear of negative evaluation (items 1, 2, 5, 6, 8, and 10); and (2) social avoidance and distress (items 3, 4, 7, and 9). Each item has three response options (0–2) that rate degree of anxiety: 0 = no anxiety symptoms; 1 = moderate anxiety symptoms, and 2 = severe anxiety symptoms. The total score of the SASC ranges from 0 to 20, with higher scores reflecting higher anxiety. A total score ≥ 8 indicates the possibility of social anxiety disorder.

#### Depression Self-rating Scale for Children (DSRSC) [17]

The DSRSC, which was developed by Birleson for children age 7–13 years, contains 18 items that are rated as no = 0, sometimes = 1, and often = 2. High scores indicate more severe depression. Items 1, 2, 4, 7, 8, 9, 11, 12, 13, and 16 are reverse scored, so that no = 2, sometimes = 1, and often = 0. The items are summed to obtain a total score, which ranges from 0 to 36. Higher scores indicate a greater degree of depression, with a total score ≥15 indicating the possibility of depressive disorder.

### Procedure

The researchers created a questionnaire for an online survey App, and distributed it to students or their guardians through social software. The students completed the GIQ, SASC, and DSRSC with the help of their guardians using a mobile phone or computer.

### Statistical Analysis

All statistical analyses were performed with SPSS (v22.0) software. Frequency count data are expressed as percentages (%), and data from the measures are expressed as mean ± standard deviation. The chi-square (χ^2^) test was used to compare frequency counts between groups, and Student’s *t*-test was used to compare the scores on the measures with their Chinese norms. We used multiviate regression and logistic regression for binary dependent variables to analyze variables associated with students’ mental state. P < 0.05 was considered statistically significant.

## Results

### Demographic characteristics and general information

A total of 1974 questionnaires were distributed and 1646 were recovered, of which 1620 questionnaires were included in the analysis after removing students who were less than 7 years old or more than 13 years old. The average age of the sample was 10.10 ± 1.63 years old, including 169 (10.4%) students in first grade, 132 (8.1%) in second grade, 305 (18.8%) in third grade, 347 (28.2%) in fourth grade, 323 in fifth grade (36.1%), and 344 (35.7%) in sixth grade. There were 835 males (52.2%) and 785 females (47.8%), all of whom are *Han* nationality, and no family members or students reported being infected.

As shown in Table 1, 25.2% of the students were only accompanied by their father and/or mother during their home quarantine, and 72.8% were also accompanied by other relatives, such as grandparents; 2% were only accompanied by relatives (e.g., grandparents), but not their parents. The average daily time of parent-child activity was 3.1h ± 3.95, and more than 95% of the students maintained a good parent-child relationship with their parents during home quarantine. On average, students studied 4.1h ± 2.39 per day and played 2.9h ± 2.40; 89.6% exercised more than 3 times a week, and 45.5% took out-of-school training classes (i.e., remote courses). In addition,91.2% of the students had a regular schedule for learning and rest. All the students understood what COVID-19 was to some extent, mainly based on family, Internet, TV, and other sources. Only a few students (8.8%) worried about being infected, but 4.9% of them experienced negative emotions, such as being irritable (47.4%), listless (42.1%), and unable to be calm and concentrate on learning (26.6%).

**Table 1:** General Information about the primary school students during home quarantine

| Items | Classification | Number of cases (proportion) |
| --- | --- | --- |
| Grade | Low grades (grades 1-3) | 606(37.4%) |
|  | High grades (grades 4-6) | 1014(62.6%) |
| Sex | Male | 835(52.2%) |
|  | Female | 785(47.8%) |
| Companionship | Only accompanied by father and / or mother | 408(25.2%) |
|  | Accompanied by parents and other relatives | 1179(72.8%) |
|  | Without parents | 33 (2.0%) |
| Parents-child relationship | harmony | 1580(97.5%) |
|  | discord | 40(2.5%) |
| Exercise per week | 3 times and above | 1451(89.6%) |
|  | Less than 3 times | 169(10.4%) |
| Extracurricular training classes | Yes | 737(45.5%) |
|  | No | 883(54.5%) |
| Regular learning and rest | Yes | 1477(91.2%) |
|  | No | 143(8.8%) |
| Understanding of COVID-19 | Yes | 1620(100%) |
| Worry about being infected | Yes | 142(8.8%) |
|  | No | 1478(91.2%) |
| Emotion changes | Positive | 1540(95.1%) |
|  | Negative | 80(4.9%) |
| Main negative emotions | Easy to be irritable | 37(47.4%) |
|  | Listless | 33(42.1%) |
|  | Unable to calm and concentrate on learning | 21 (26.6%) |

### Social anxiety

The mean total SASC score of the 1620 students was 3.90 ± 3.73, including 2.53 ± 2.63 for fear of negative evaluation and 1.37 ± 1.62 for social avoidance and distress. We compared their mean scores with the SACS norm scores of children from urban areas of 14 large and medium-sized cities in China, which were developed by a National Collaborative Group of the SASC in 2006 [18]. We found the scores for both dimensions and the total score of our study participants were significantly higher than the norm scores (Table 2).

**Table 2:** SASC scores of the primary school students during home quarantine

|  |  | fear of negative<br>evaluation | social avoidance<br>distress | Total score |
| --- | --- | --- | --- | --- |
| Overall | Participants (N=1620) | 2.53±2.63 | 1.37±1.62 | 3.90±3.73 |
|  | Norm (N=2019) | 2.24±2.5 | 1.24±1.5 | 3.48±3.47 |
|  | P-value | <0.01 | <0.05 | <0.01 |
| Female | Participants (n=785) | 2.75±2.68 | 1.47±1.67 | 4.22±3.83 |
|  | Norm (n=692) | 2.05±2.37 | 1.19±1.47 | 3.24±3.27 |
|  | P-value | <0.01 | <0.01 | <0.01 |
| Male | Participants (n=835) | 2.31±2.57 | 1.28±1.57 | 3.59±3.59 |
|  | Norm (n=677) | 1.89±1.89 | 1.26±1.56 | 3.19±3.28 |
|  | P-value | <0.01 | >0.05 | <0.05 |
| Age | Low grades (n=607) | 2.55±2.54 | 1.49±1.63 | 4.04±3.65 |
|  | High grades (n=1013) | 2.51±2.68 | 1.31±1.61 | 3.82±3.77 |
|  | P-value | >0.05 | <0.05 | >0.05 |
| Social anxiety | Positive (n=279) | 6.67±2.10 | 3.74±1.66 | 10.41±2.59 |
|  | Negative (n=1341) | 1.66±1.77 | 0.88±1.09 | 2.54±2.17 |
|  | P-value | <0.01 | <0.01 | <0.01 |

A total of 279 students with social anxiety were identified, for a detection rate of 17.2%. The mean score for these students was 10.41 ± 2.59, ranging from 8 to 20. The mean score was 6.67 ± 2.10 for negative evaluation, and 3.74 ± 1.66 for social avoidance and distress, which were much higher than the scores of the students who did not have social anxiety (P < 0.01) (Table 2).

Female students had significantly higher SASC scores than the norm scores (P < 0.01) on all three measures (Table 2). Male students had significant higher scores on the fear of negative evaluation dimension than the norm scores (P < 0.01), but there was no significant difference between their scores for social avoidance and distress and the norm scores. When we divided the students into two groups – low grades (grades 1–3) and high grades (grades 4–6) – we found that the mean score for social avoidance and distress was significantly larger in the low grades than the high grades (P < 0.05).

### Depression

Our survey found that the mean total score of the DSRSC during home quarantine was 5.67 ± 4.97 (range = 0–33), which was much lower than the norm score of the DSRSC for children from urban areas in China (9.84 ± 4.73) (P < 0.05) [19]. The scores of all the positive items were significantly lower than the norms, and the scores for most negative items were significantly lower than the norms (Table 3). Our results indicate that primary school students who were quarantined at home were less likely to be depressed than the norm.

**Table 3:** DSRSC scores of the primary school students during home quarantine

| Items | Participants<br>(N=1620) | Norm<br>(N=1943) | P-value |
| --- | --- | --- | --- |
| 1.I look forward to things as much as I used to | 0.39±0.16 | 0.80±0.49 | 0.000 |
| 2.I sleep very well | 0.27±0.09 | 0.52±0.24 | 0.000 |
| 3.I feel like crying | 0.32±0.57 | 0.32±0.58 | 0.500 |
| 4.I like to go out to play | 0.48±0.21 | 0.74±0.43 | 0.000 |
| 5.I feel like running away | 0.10±0.35 | 0.12±0.36 | 0.095 |
| 6.I get tummy aches | 0.30±0.50 | 0.43±0.58 | 0.000 |
| 7.I have lots of energy | 0.22±0.06 | 0.82±0.53 | 0.000 |
| 8.I enjoy my food | 0.25±0.07 | 0.70±0.38 | 0.000 |
| 9.I can stick up for myself | 0.30±0.09 | 0.66±0.35 | 0.000 |
| 10.I think life isn't worth living | 0.19±0.46 | 0.25±0.52 | 0.000 |
| 11.I am good at things I do | 0.79±0.40 | 1.12±0.84 | 0.000 |
| 12.I enjoy the things I do as much as I used to | 0.32±0.10 | 0.80±0.49 | 0.000 |
| 13.I like talking with my family | 0.37±0.13 | 0.72±0.43 | 0.000 |
| 14.I have horrible dreams | 0.42±0.52 | 0.42±0.59 | 0.500 |
| 15.I feel very lonely | 0.26±0.52 | 0.34±0.58 | 0.000 |
| 16.I am easily cheered up | 0.24±0.07 | 0.59±0.31 | 0.000 |
| 17.I feel so sad I can hardly stand it | 0.18±0.42 | 0.26±0.52 | 0.000 |
| 18. I feel very bored | 0.30±0.52 | 0.47±0.63 | 0.000 |

A total of 102 students with depression were identified, for a detection rate of 6.3%. Their mean total DSRSC score was 18.96 ± 3.89 (range = 15–33), which was much higher than the mean score of the students without depression (4.78 ± 4.56) (P < 0.01). The mean DSRSC score of the high-grade group (grades 4–6) was 6.04 ± 5.28, which was significantly higher than those of the low-grade group (grades 1–3) (5.06 ±4.33) (P < 0.01). No significant difference in scores was found between the male and female students.

### Risk factors for social anxiety and depression

Our results showed that students with social anxiety were accompanied by fewer relatives than those without social anxiety (P < 0.05) (Table 4). We also found that students with depression were accompanied by fewer relatives than those without depression (P < 0.05) (Table 4), especially those who were accompanied by both their mother and father (P < 0.01). In addition, students with social anxiety or depressive disorder were significantly more likely to have a poor parent-child relationship, deterioration of the parent-child relationship, increased conflicts with parents, irregular learning and rest, less physical activity, more worry about being infected, and negative mood (P < 0.05) (Table 4). Further investigation found that conflicts with parents mainly included the study time schedule (67.2%), the use of electronic products (58.4%), and rest time (42.2%).

**Table 4:** Risk factors for social anxiety and depressive disorders durin home quarantine

| Variables | Social anxiety |  | P-value | Depression |  | P-value |
| --- | --- | --- | --- | --- | --- | --- |
|  | Positive | Negative |  | Positive | Negative |  |
|  | (n=279) | (n=1341) |  | (n=102) | (n=1518) |  |
| Number of relatives accompanied | 3.34±1.35 | 3.53±1.32 | 0.0295 | 3.22±1.32 | 3.52±1.33 | 0.028 |
| Parents accompanied together | 270 (86.0%) | 1187 (88.5%) | 0.242 | 78 (76.5%) | 1349 (88.9%) | 0.000 |
| Parent-child activity time per day | 3.03±3.53 | 3.34±4.02 | 0.232 | 2.62±4.10 | 3.34±3.93 | 0.074 |
| Poor parent-child relationship | 18 (6.5%) | 22 (1.6%) | 0.000 | 21 (20.6%) | 19 (1.3%) | 0.000 |
| Deterioration of parent-child relationship | 38 (13.6%) | 58 (4.4%) | 0.000 | 38 (37.3%) | 58 (3.8%) | 0.000 |
| Increased conflicts with parents | 119 (42.6%) | 320 (23.8%) | 0.000 | 57 (55.8%) | 382 (25.2%) | 0.000 |
| Irregular work and rest | 38 (13.6%) | 104 (7.8%) | 0.002 | 23 (22.6%) | 119 (7.9%) | 0.000 |
| Less physical activity | 42 (15.1%) | 126 (9.4%) | 0.005 | 17 (16.7%) | 151 (10.0%) | 0.031 |
| Gaining weight | 110 (39.4%) | 457 (34.1%) | 0.088 | 37 (36.3%) | 530 (34.9%) | 0.780 |
| Learning time per day | 4.06±2.32 | 4.07±2.40 | 0.949 | 4.26±2.62 | 4.06±2.37 | 0.413 |
| Entertainment time per day | 2.84±1.74 | 2.90±2.51 | 0.703 | 2.78±1.98 | 2.90±2.42 | 0.624 |
| Extracurricular training classes | 148 (53%) | 666 (49.7%) | 0.304 | 53 (52%) | 761 (50.1%) | 0.721 |
| Worry about being infected | 55 (19.7%) | 155 (8.6%) | 0.000 | 19 (18.6%) | 51 (9.9%) | 0.006 |
| Negative mood | 39 (14%) | 58 (4.3%) | 0.000 | 24 (23.5%) | 73 (4.8%) | 0.000 |

We conducted a logistic regression with social anxiety (yes = 1, no = 0) as the dependent variable, and the following set of independent variables: sex, age, number of relatives accompanying students during quarantine, whether parents accompanied them together, parent-child activity time per day, the parent-child relationship, changes in the parent-child relationship during home quarantine, contradictions and conflicts, physical activity, regular work and rest, weight gain, extracurricular training classes, worry about being infected, emotional changes, learning time per day, and entertainment time per day as independent variables (for the assignment table, see Supplemental Table 2). We found that being male, deterioration of the parent-child relationship, increased conflicts with parents, irregular learning and rest, worrying about being infected, and negative mood were risk factors for social anxiety (Table 5). Students with a deteriorating parent-child relationship were 2.068 times more likely to suffer from social anxiety than students with no change in parent-child relationship during the home quarantine. Students who worried about being infected were 2.206 times more likely to suffer from social anxiety than those who did not worry about being infected. And students with negative emotional changes were 2.438 times more likely to have social anxiety than those with positive emotional changes (Table 5).

**Table 5:** Logistic regression of risk factors for social anxiety and depression during home quarantine

| Risk factors | Social anxiety |  |  | Depression |  |  |
| --- | --- | --- | --- | --- | --- | --- |
|  | Wald | OR-value | P-value | Wald | OR-value | P-value |
| Sex | 8.866 | .663 | .003 | 1.473 | .755 | .225 |
| Age | 3.445 | .923 | .063 | 3.945 | 1.169 | .047 |
| Number of relatives accompanying | 2.853 | .912 | .091 | .915 | .915 | .339 |
| Parents accompanied together | .011 | .978 | .915 | 3.392 | .578 | .066 |
| Parent-child activity time per day | .916 | .973 | .338 | 2.930 | .905 | .087 |
| Parent-child relationship | 1.391 | 1.613 | .238 | 9.366 | 3.697 | .002 |
| Changes in parent-child relationship | 7.049 | 2.068 | .008 | 31.906 | 5.744 | .000 |
| Increased conflicts with parents | 19.917 | 1.975 | .000 | 2.210 | 1.510 | .137 |
| Physical activity | 1.498 | 1.239 | .221 | 8.094 | 2.070 | .004 |
| Irregular learning and rest | 6.126 | 1.327 | .013 | 4.011 | 1.439 | .045 |
| Weight gain | 2.529 | 1.253 | .112 | .188 | .900 | .664 |
| Extracurricular training classes | .961 | 1.146 | .327 | .052 | .948 | .820 |
| Worry about being infected | 17.428 | 2.206 | .000 | .644 | 1.292 | .422 |
| Emotional changes | 13.819 | 2.438 | .000 | 9.957 | 2.726 | .002 |
| Entertainment time per day | .035 | .994 | .852 | .044 | .987 | .833 |
| Learning time per day | .068 | 1.008 | .794 | .255 | 1.023 | .614 |
| Constant | 1.404 | .494 | .236 | 16.598 | .015 | .000 |

We conducted logistic regression with depression (yes = 1, no = 0) as the dependent variable, and the same independent variables used in the analysis of social anxiety (Supplemental Table 2) for binomial logistic regression analysis. We found that older age, a poor parent-child relationship, deterioration of the parent-child relationship, less physical activity, irregular work and rest, and negative mood were risk factors for depression. Students with a poor parent-child relationship were 3.697 times more likely to suffer from depression than students with a good parent-child relationship. Students with a deteriorating parent-child relationship were 5.744 times more likely to suffer from depression than were those with no change in their parent-child relationship. Students with negative emotional changes during quarantine were 2.438 times more likely to have depression than those with positive emotional changes during quarantine (Table 5).

## Discussion

China has closed schools and adopted home quarantine measures since the outbreak of COVID-19 to control the spread of SARS-CoV 2 [3]. Our research found that primary school students, overall, were susceptible to social anxiety during home quarantine. We found 279 out of 1620 (17.2%) primary school students had social anxiety. Compared with their norms, primary school students who were quarantined at home had significantly higher social anxiety scores, indicating that the prolonged (at least one month) period of strict home quarantine during the epidemic tended to increase social anxiety, consistent with previous research on the social anxiety of children after major disasters [20]. Communities, families and schools should pay more attention to the social anxiety status of primary school students during the epidemic. Our study found that males were more likely than females to have social anxiety during the epidemic, which differs from the findings of previous studies that investigated children living a normal life [21,22]. The children in our study were quarantined at home during the epidemic and the active nature of these students, especially the male students, could not be fully expressed in the relatively small indoor spaces.

The psychological problems of the 279 students with social anxiety can be partially attributed to the deterioration of their parent-child relationship during home quarantine, and increased conflict between parents and children. Students with social anxiety were more likely to have a deteriorated parent-child relationship (13.6% vs 4.4%) and conflicts with parents (42.6% vs 23.8%). Social anxiety is a common response of primary school students to a deterioration in the parent-child relationship and conflict [23]. Results from twins and family studies over the past three decades have confirmed that childhood social anxiety is influenced by both genetic factors [24], and parental factors, including parenting styles and parent-child interactions [25]. The deterioration of the parent-child relationship is usually caused by the limited or inconsistent expression of care and warmth, and excessive protection and control [25]. Family interventions should be taken based on the five basic principles of promoting a sense of security: helping children learn to be calm and stable in a crisis and have a sense of self-worth and a sense of family or group belonging, promoting social ties, and instilling hope [26].

Irregular learning and rest is another risk factor for social anxiety in primary school students, as this leads to irregular diet, and sleep and physical activity, resulting in a decline in sleep quality. A 2018 study [27] reported that poor sleep quality was related to increased anxiety. In addition, regular aerobic exercise can help reduce anxiety [28]. Keeping a regular life schedule, such as learning and rest time during home quarantine, can prevent or decrease social anxiety among primary school students. In addition, worry about being infected and negative emotions during the epidemic could have increased the likelihood of social anxiety. Since COVID-19 is a novel infectious disease without specific drugs and vaccines [3], the wide and rapid spread of the epidemic during its early stage could have easily caused panic in the population and led to psychological distress [3]. Scientific education about COVID-19, especially preventive and protective measures, may help ease fear of infection and reduce anxiety.

Interestingly, we found the level of depressive symptoms of the students quarantined at home was significantly lower than the norm, which is inconsistent with previous studies on depression among children facing major disasters [29,30]. The reason for this may be that even though the COVID-19 epidemic was widespread, no students or family members in our study reported being infected. They were quarantined at home, which was a safe place during the early stage, and had not faced the crisis directly. More importantly, compared to ordinary times, home quarantine increased the time for parent-child interaction, especially time with parents, as the students’ parents were also quarantined at home. However, no significant difference was found in parent-child activity time between students with and without depression in our study. The increased frequency and quality of interactions between parents and children can improve children’s depression [31,32]. Moreover, the increased parent-child interaction time can reinforce a child’s attachment to and approval from the family, and depression is inversely related to attachment, family support, and approval [33].

We found the students with depressive disorders had a poor parent-child relationship, which was a risk factor for depression during home quarantine. Family dysfunction often persists when the relationship between parents and children deteriorates [34], and leads to chronic and persistent relapses of depression [35]. Therefore, it is very important to improve parent-child relationships during home quarantine. We also found that less physical activity and irregular learning and rest schedules were risk factors for depression. Students who exercised more than three times a week had a significantly lower risk of developing depressive symptoms. A recent study conducted by Kandola et al. found that increased sedentary behavior and less activity were associated with an increased risk of depressive symptoms at the age of 18 [5], which is consistent with our results. In addition, negative emotions of the students related to the virus outbreak can also lead to depression. The thickness of the frontal cortex of patients with depression is different from that of healthy people [36], and negative emotions are usually found to be related to frontal dysfunction [37]. A loving and comforting family environment can be helpful for reducing negative emotions.

## Conclusion

Our study indicates that primary school students quarantined at home during the COVID-19 epidemic were prone to social anxiety but less likely to suffer from depression. Although the two psychological disorders share common risk factors (e.g., deteriorating parent-child relationship, increased conflicts with parents, and irregular learning and rest), we believe that social isolation itself may be the main cause of social anxiety, whereas depression is affected more by parent-child relationships and interactions. Close attention should be paid to the psychological status of children and adolescents during a virus outbreak, especially those who are quarantined at home. Early identification of and interventions for social anxiety and depression should be considered when developing strategies and countermeasures against COVID-19 epidemics for schools and communities.

## Data Availability

The data that were analyzed to produce the findings of this study are available from the corresponding author upon request

## Acknowledgments

We thank all the participants and their guardians for the agreements with our questionnaire survey.

## Author contributions

YZ, JL, and MZ contributed equally to this paper. JL, ZC and CJ contributed to the study conceptualization and design. MZ, BJ, XL, and ZC contributed to data collection, and YZ, ZC, NW, and CJ contributed to data analysis. YZ, JL, and CJ participated in data interpretation and manuscript preparation. YZ, JL, MZ, and CJ did the literature search. All authors read and approved the final version. The corresponding authors attest that all listed authors meet authorship criteria and that no others meeting the criteria have been omitted. CJ is the guarantor.

